# Statistical Inference for Coronavirus Infected Patients in Wuhan

**DOI:** 10.1101/2020.02.10.20021774

**Authors:** Yong-Dao Zhou, Jianghu James Dong

## Abstract

**Importance:** The new coronavirus outbreak has seriously affected the quality of life in China. Wuhan is the disaster area, where the number of cases has increased rapidly. However, the current measures of infected patients in Wuhan are still underestimated.

**Objective:** To estimate the overall infected patients in Wuhan from several sampled data. The correct estimated infected patients can be helpful for the government to arrange the needed beds in hospital wards to meet the actual needs.

**Design:** We proposed to use the sampling survey to estimate the overall infected patients in Wuhan. The sampling survey is a kind of non-comprehensive survey. It selected some units from all the survey objects to carry out the survey and made the estimation and inference to all the survey objects. Sampling surveys can obtain information that reflects the overall situation, although it is not a comprehensive survey.

**Setting:** We estimated the overall infection rate in Wenzhou city, which has a better data collection system. Simultaneously, another different samples of Wuhan tourists to Singapore will be used to validate the infection rate in Wenzhou city. Combined these two samples, we give the estimation of the number of infected patients in Wuhan and other prefecture-level cities in Hubei Province.

**Participants:** The number of people who returned from Wuhan to Wenzhou was selected from the daily notification of the pneumonia epidemic caused by a new coronavirus infection in the city.

**Exposures for observational studies:** The daily rate of the pneumonia epidemic caused by the new coronavirus infection in Wenzhou City. The numerator is the number of people diagnosed and whether each person diagnosed had a history of living in Wuhan. The denominator is the total number of people returning to Wenzhou from Wuhan. Based on this rate, it is reasonable to predict the number of the infected patients.

**Main Outcome(s) and Measure(s):** According to the most conservative estimate from our proposed sampling method, at least total 54,000 infected patients are in Wuhan. Therefore, the current 8,000 beds in hospital wards and the 20,000 beds in square-class hospitals are far away from meeting the actual needs.

## Methods

### Study Design and Participants

Total of 180,000 Wenzhou people was in business/school or worked in Wuhan. Wenzhou city had detected 33,000 people returning from Wuhan and its surrounding areas from the news conference of the coronavirus outbreak by Wenzhou city government on Jan 29, 2020. Among these 33,000 people, the number of cases of pneumonia infected by the new coronavirus in Wenzhou city was as follows:

According to Table 1, the total number of confirmed cases in Wenzhou was 448. The number of people who suffered from Wuhan and its surrounding areas was 202, and the number of people who returned to Wenzhou from Wuhan and its surrounding areas was 33,000; Therefore, the infection rate of the people returning to Wenzhou from Wuhan is about 0.61%. It is important to note that all these people returned before January 29, 2020. This indicates that the infection rate was about 0.61% by January 29, 2020.

**Table 1:** Distribution of the new coronavirus in Wenzhou city

| Table 1: Distribution of the new coronavirus in Wenzhou city |  |  |
| --- | --- | --- |
| Time | Infected patients | Patients who traveled from Wuhan |
| Jan. 27 | 60 | 57 |
| Jan. 28 | 54 | 39 |
| Jan. 29 | 58 | 36 |
| Jan. 30 | 55 | 34 |
| Jan.31 | 14 | 3 |
| Feb.1 | 24 | 6 |
| Feb.2 | 26 | 3 |
| Feb.3 | 49 | 11 |
| Feb.4 | 24 | 3 |
| Feb.5 | 32 | 4 |
| Feb.6 | 25 | 3 |
| Feb.7 | 17 | 2 |
| Feb.8 | 10 | 1 |
| Feb.9 | 16 | 0 |
| Total | 448 | 202 |

### Statistical sampling method

From the statistical sampling, Wenzhou people who were in business/school or worked in Wuhan can be used as a sample of Wuhan residents. The infection rate of the sample can be used to estimate the overall infection rate. Due to the impact of the Spring Festival and the epidemic situation, more than 5 million people have left Wuhan from the news conference of the coronavirus outbreak by Wuhan Government on Jan 26, 2019. Nine million more residences remained in Wuhan. This shows that the population of Wuhan is 14 million. From sampling, the infection rate of the sample is a good estimate of the overall infection rate.

However, given the number of samples of the person returning Wenzhou, the rate of total 14 million was less than 0.24%. Estimates of infection rates may be inaccurate, possibly because many people from Wenzhou were doing business in Wuhan and were more likely to contact people than the others from other places in Wuhan. Statistically, the sample of samples may not represent the overall situation well. Therefore, additional data are required to verify each other and correct infection rates. To this end, total 10,680 Wuhan tourists to Singapore from December 30, 2019 to January 22, 2020 from government reports are included as a subsample. As of February 7, 2020, out-of-town travelers were diagnosed in Singapore, while some tourists did not suffer from the disease during their stay in Singapore but returned to China or went to a third country to suffer from the disease. For example, on January 25, 2020, three Malaysian diagnosed patients arrived in Malaysia two days after they arrived in Singapore. On February 7, the Ministry of Health of Malaysia announced the new diagnosis of a 59-year-old Chinese woman who entered Malaysia from Singapore on January 21, 2020; In addition, 335 passengers arrived at Hangzhou airport from Singapore on January 24, 2020. Among the 335 passengers on board, there were 116 passengers from Wuhan. As of February 5, 2020, a total of nine passengers on board were confirmed. In all, at least 33 of the 10,680 people were diagnosed. The infection rate shall not be lower than 0.3%. It should be noted that this infection rate referred to the infection rate before January 23. By January 29, 2020, the infection rate would be higher. The infection rate in Wuhan was estimated at 0.3-0.6% as of January 29 after combined with the sampling data Wuhan residents from Wenzhou and the sampling data from Wuhan tourists to Singapore.

## Results

Wuhan city had about 42,000 people infected by Jan 29, 2020, which was calculated based on the 0.3% rate. 27,000 people were remaining in Wuhan, while 15,000 people were living in non-Wuhan areas. If based on the 0.6% rate, Wuhan city had about 84,000 infected people. 54,000 people were remaining in Wuhan, while 30,000 people were living in non-Wuhan areas

The number of confirmed cases in Wenzhou has more than doubled under strict preventive measures such as the collection of suspected cases and concentrated isolation of those in close contact during passed ten more days since January 29, 2020. However, Wuhan City isolated suspected cases and diagnosed patients with mild illness in their homes, which increased the possibility of family infection during this period. Therefore, the number of people infected in Wuhan was estimated to be between 54,000 and 90,000. The number of people infected in other areas was estimated at 30-50,000. The total number of people infected nationwide is 84,000-140,000. Since 70% of the 5 million people who left Wuhan were returning to other parts of Hubei, the estimated number of people infected in the non-Wuhan region was 21,000-35,000. The number of infected persons in other provinces was estimated at 9-15,000.

Total of 10,540 people had been diagnosed in other provinces on February 9, 2020. This data is accurate because it was not intentionally not subjected to nucleic acid testing. Combined with the number of infected and undiagnosed patients, the number of infected persons in other provinces fell within the range of 9-15,000, which is more consistent with the estimated value of this paper. The number of people returned from Wuhan to other provinces is 1.5 million, accounting for more than 10% of the total population of 14 million. Thus, the infection rate of 0.3% is a very conservative rate before January 29, 2020.

## Conclusions

(1) The current measures of infected patients are yet to be upgraded. According to the most conservative estimate of 54,000 infections in Wuhan. The current 8,000-plus beds in hospital wards and the 20000 beds in square-class hospitals are far away from meeting the actual needs.

(2) According to the most conservative estimate of 21,000 infections in the cities in Hubei province except Wuhan, the number of suffered people, 12729, by February 9 2020, are also to be upgraded.

